# Use of Machine Learning for Long Term Planning and Cost Minimization in Healthcare Management

**DOI:** 10.1101/2021.10.06.21264654

**Authors:** Sadia Binta Kabir, Salman Sadiq Shuvo, Helal Uddin Ahmed

## Abstract

The Healthcare system of a country is a crucial infrastructure that requires long-term capacity planning. The covid 19 outbreak pointed to the necessity of adequate hospital capacity, especially for developing countries like Bangladesh. The existing infrastructure planning of these countries emphasizes short-term goals and lacks vision planning for a long time horizon. It is in the country’s best interest to make long-term capacity expansion plans, a strategy the developed countries banked to provide adequate healthcare facilities to their residents. However, no single solution is appropriate for a different region. Hence, it is required to comprehensively study the situation and constraints of the specific region before providing expensive capacity expansion plans. This work focuses on applying a deep Reinforcement Learning based long-term hospital bed capacity expansion plan. We utilize the RNN-LSTM based population forecast, deep Reinforcement Learning (RL) based policy-making, and state-of-the-art Artificial Intelligence techniques to provide a solution. We perform a case study for the Abhaynagar Upazila of Jessore, one of the largest cities in the southwest part of Bangladesh, to analyze the benefits of such an approach compared to existing myopic policies. The experiment results show that the deep RL-based policy significantly minimizes cost over a 30-year expansion plan.

## I. Introduction

### A. Motivation

Hospital bed shortage is a pervasive problem for developing countries. This problem is much more severe in populated countries like Bangladesh. The hospital bed crisis makes people suffer more in the rural and district area, where people depend solely on government hospitals. The inconsistency in the number of patients in a hospital ward makes it challenging to estimate the required bed for the patients. The number of patients requiring overnight stay higher than the hospital’s capacity may lead to overflow and denial of service to critical patients. Hospital capacity augmentation requires more doctors, additional space, medical equipment, furniture, and nursing facilities that incur high costs for the administration. Though observing the highest needs of beds and provide accordingly seems to be a solution. However, that is very much impractical to a developing country like Bangladesh. Thus it is a significant concern of planning for the authority.

Bangladesh has made some improvements in the health care system; however, with these improvements, people’s access to quality healthcare becomes insignificant due to the lack of caregiving infrastructure such as hospital beds. When large populations and remote geographic locations come to the scenario, the inadequate healthcare system grows more vulnerable to unexpected high demands during pandemics or disasters. World Bank reports that there are only 0.8 hospital beds in Bangladesh per 1000 people, compared to the global mean of 2.7. This scarcity has made getting minimum treatment highly unlikely and sometimes encourages corruption. On the other hand, a sudden rise in hospital bed demand during the pandemic or natural calamities causing significant damage to the overall public health in Bangladesh. All of these urges the necessity of well-educated future development plans for hospital capacities. There are unofficial costs, including bribes for getting the hospital cabin or beds. It shows that ordinary people have to spend ten to fifteen times more than the official cost of services.

Bangladesh has the required capacity to offer preventive and promotional healthcare to people all over the country. Nevertheless, many people, especially in rural areas, have insufficient access to healthcare facilities. The rural areas of Bangladesh account for nearly 63% of the country. The government-owned hospital generally serves the rural population. Thus the health authority of the region is solely responsible for accommodating the hospitalization of the residents. So a sustainable skeleton to forecast future hospital bed demands comprises a significant part of future up-gradation plans of the hospitals. In this work, We consider a rural setup with health administration as the decision-maker (i.e., RL agent) and hospital capacity and population of the region as the agent’s environment. This paper aims to provide a reinforcement learning-based optimal policy to plan for future hospital bed augmentation.

### B. Literature Review

Many pieces of research are done on hospital occupancy [1], [2], but most of these are for short time projection which generally projects ICU bed requirement [1]. Extended terms are done for commercial or private sector use [2]. For general hospital admission prediction, The work provides a guideline for the expected number in hospital occupancy levels to include and deal with the variance [3]. This paper provides a foundation for our policymaking for Bangladesh. Further, The work [4] provides an Operation Theatre (OT) capacity policy from an economic point of view. The cost-benefit analysis is fundamental for any long-term capacity expansion planning that we accommodate in this work. The work [5] insists on the importance of hospital bed-occupancy rates in assessing the success of a health care facility to serve its patients.

Many works follow data-driven methods and machine learning (ML) techniques for predictive tasks [6]. The works in [7], [8] utilize feed-forward Neural Network and Recursive Neural Network (RNN) based hospital bed occupancy forecast. Besides supervised and unsupervised learning, reinforcement learning (RL), a central area of ML, is an efficient technique for generating agent-based scenarios for planning resource optimization. Many works have utilized this technique for natural disaster management [9], transportation systems [10], electrical systems [11], and health sector [12], [13].

Recently, the work [14] has discussed the application of ML-based hospital up-gradation policy for Bangladesh. However, they run their experiments for a hypothetical rural Bangladesh region, thus lacking a practical zone’s feasibility analysis. Furthermore, the model is backed up by an accurate population forecast, which is not pragmatic for many rural parts of Bangladesh. The work provided an efficient guideline for this problem; however, it lacks some tools for practical implementation. So, we intend to address the following short-comings of the paper [14]:

1. The work uses world bank data for the population growth of the whole of Bangladesh. An oversimplification that we address by focusing the experiments for a particular Upazila of Jessore district.
2. The work hypothesize that the authority has a population forecast for the region; however, the population growth dynamic for a small region is difficult to predict. RNN-LSTM based forecasting methods follow the trend of time-series data, hence can adapt to capture the trend of population growth for a region.

### C. Contributions

The contributions are:

1. Simulation tool to predict future demand of hospital beds in Abhaynagar Upazila of Jessore.
2. Markov Decision Process (MDP) model to optimize hospital bed augmentation policy.
3. RNN-LSTM based population forecast to support the MDP model.
4. Comprehensive evaluation to demonstrate the effectiveness of the proposed model

## II. Background

### A. Machine Learning

Machine learning is an evolution of artificial intelligence that enables self-learning and identify patterns from data and then applies that learning without human intervention. It is the task of making computers more intelligent without explicitly teaching them how to behave.

Machine learning is being used in various fields and industries. Medical diagnosis, image processing, prediction, classification, learning association, and regression are a few ML examples.

Machine learning is a similar way to how humans think, learn and improve upon past experiences. It explores data and recognizes patterns so that less human interference is required. The more data available, the more a solution with machine learning fits there so comfortably. The diagram below will explain how Machine Learning works more efficiently.

### B. Reinforcement Learning (RL)

Reinforcement learning is a training method of machine learning based on a rewarding system, where desired behavior gets the most reward. RL agent can perceive and interpret its environment, take actions and learn through trial and error. In supervised machine learning, the training data set has the answer key, and the data set trains the model. In contrast, there is no answer in reinforcement learning. However, the RL agent decides how to perform the provided task. As the data set is absent, it is required to learn from experience.

### C. Markov Decision Process (MDP)

Markov decision process is a discrete-time random control process. In situations where outcomes are partially stochastic and partially in control of decision-makers, MDP provides a framework for decision-making.

Markov decision process is in some state at each time step, and the decision-maker can choose any action from those states. After the first step, the process responds at the next step by moving into a random new state and providing the decision-maker with some corresponding reward. The chosen action determines the probability of moving the process to the next new state.

### D. Recursive Neural Network (RNN)

Recursive Neural Network (RNN) is suitable for time series prediction tasks like machine translation, speech generation, etc. From the Neural Network point of view, it specialization lies in using the previous outputs for the next output prediction.

### E. Long short-term memory (LSTM)

Long short-term memory (LSTM) is a particular type of RNN trained using backpropagation through time and overcame the main problem of RNN, the vanishing gradient [18]. Because of its use to create large recurrent networks, it can be used to address complex sequence problems in machine learning and achieve state-of-the-art results. Instead of neurons and layers, LSTM networks use memory blocks that are connected through layers. These blocks contain gates that manage the block’s state and output. A block operates upon an input sequence, and each Gate within a block uses the sigmoid activation units to control whether they are triggered or not, making the change of state and addition of information flowing through the block conditional. There are three types of gates within a unit:

Forget Gate: logically decides what information to eliminate from the block.

Input Gate: logically determines which values from the input to update the memory state.

Output Gate: logically decides what to output based on input and the memory of the block.

The overall architecture is shown in Fig. 5

**Fig. 1.**
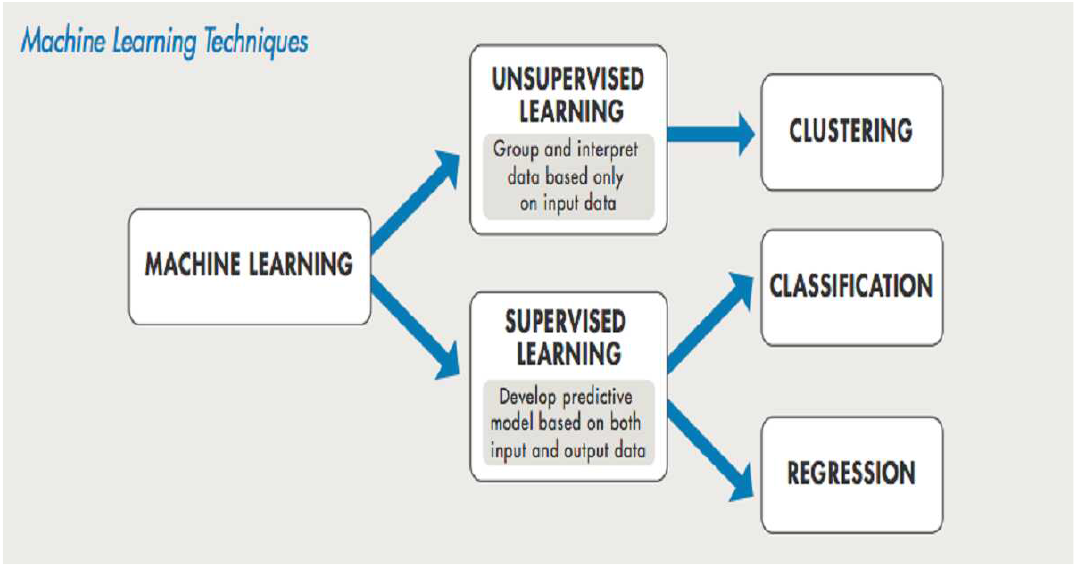
How Machine Learning works [15].

**Fig. 2.**
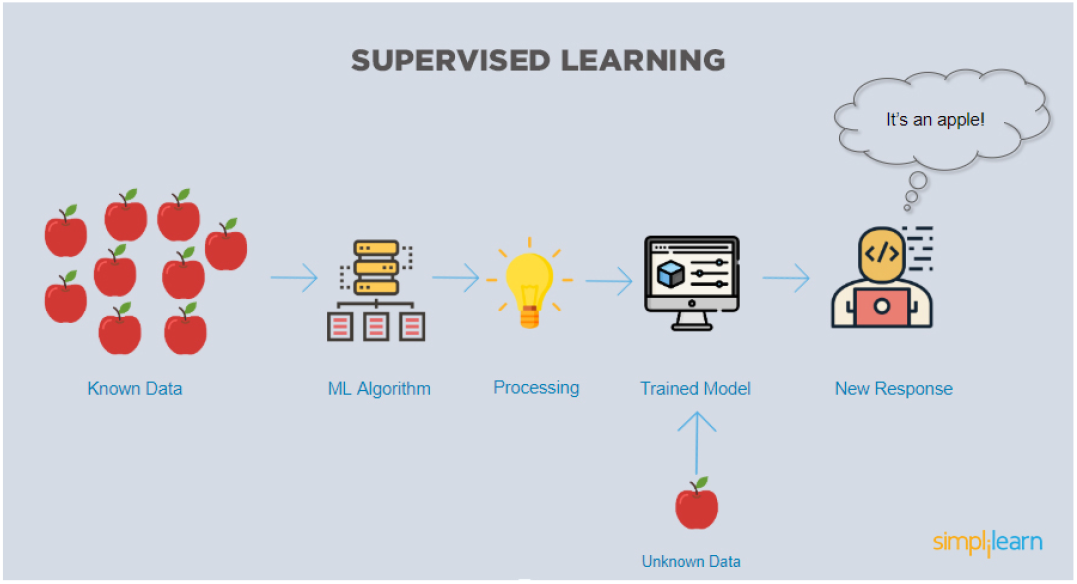
Supervised Learning [16].

**Fig. 3.**
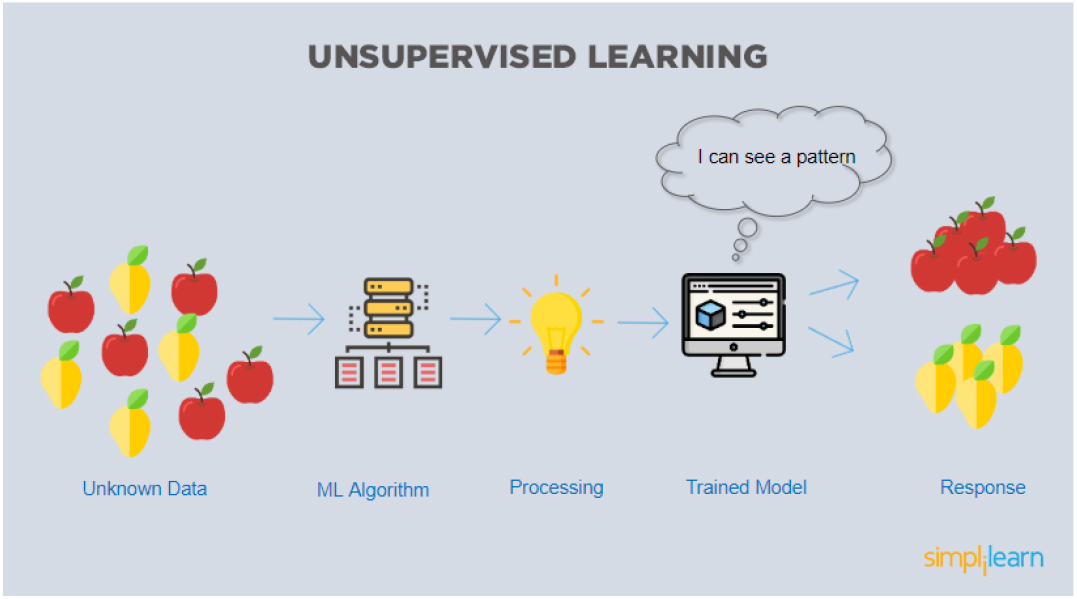
Unsupervised Learning [16].

**Fig. 4.**
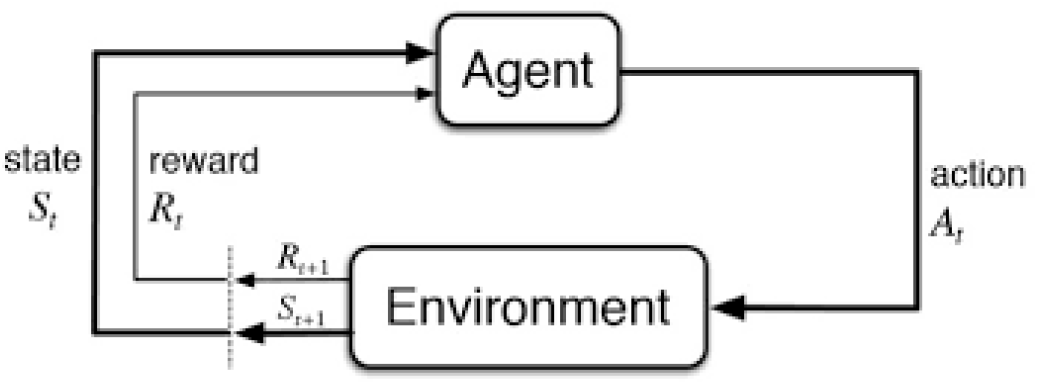
Reinforcement Learning [17].

**Fig. 5.**
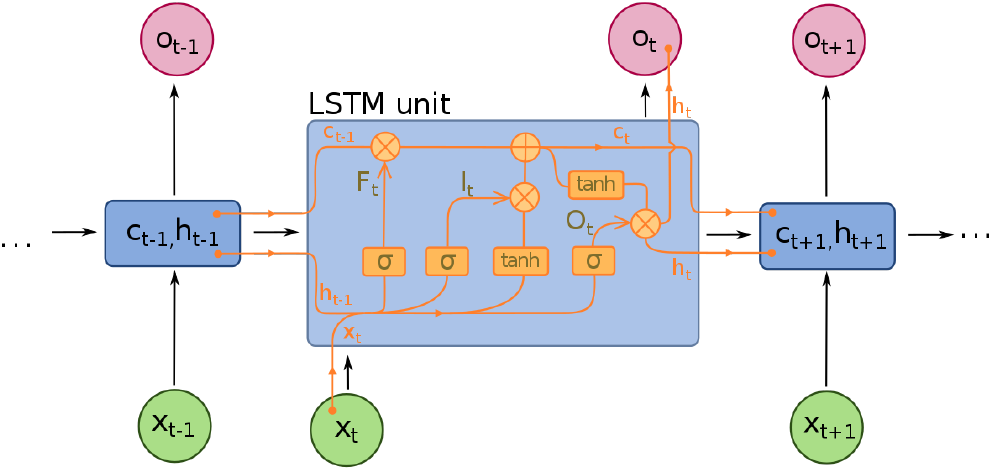
Long short-term memory (LSTM) [19].

## III. Model Developement

We design the project by the MDP model for policymaking and LSTM network for population prediction.

### A. MDP Model for Policymaking

Markov decision processes (MDP) are helpful when it is a matter of providing theoretical structure for making agent-based scenarios. We follow the MDP model provided in [14] with our modification of using an LSTM network to predict population growth. For clarity, We have explained the model in full here. The MDP agent communicates with the surroundings by taking action and receiving a reward/-cost from the situation in return. The agent’s objective is to maximize/minimize the outcomes over time by choosing optimal actions from a list of available activities. Any time, the system progresses to the next state based on the agent’s action according to a probability distribution (model-based) or a trial and error (model-free). The optimal policy decides which action to take in which state by mapping system states to actions. It can be found using dynamic programming or learned from experience employing reinforcement learning (RL), also known as approximate dynamic programming. The Advantage Actor-Critic(A2C), being a model-free RL algorithm, utilizes two neural networks to calculate the worthiness of each action for a given environment and is proper for a problem where the input elements are numerous and continuous.

Healthcare administration is the MDP agent in our model, whose action *x*_*n*_ to expand the number of beds changes the system state. All the hospitals in the Jessore district are supervised and regulated by the government healthcare authority. For generalization, we consider them as a single entity. If the number of patients admitted to a hospital concurrently is higher than the capability, some hospitals will be declined the bedding facility and thus treatment. This situation creates difficulty for the patients and causes costs to the agent, which can be a rare but significant event for a fair-sized hospital. On the other hand, if the hospital size is inadequate, this event can become commonplace, even regular. An ever-growing population of Jessore district is another factor that favors the need to expand hospital capacity. The MDP agent’s objective is to select additional beds at each decision time(e.g., every year). Growing the capacity yields cost; therefore, the agent looks for an optimal amount to increase its capacity.

The components of the MDP model shown in Fig. 6 are:

**Fig. 6.**
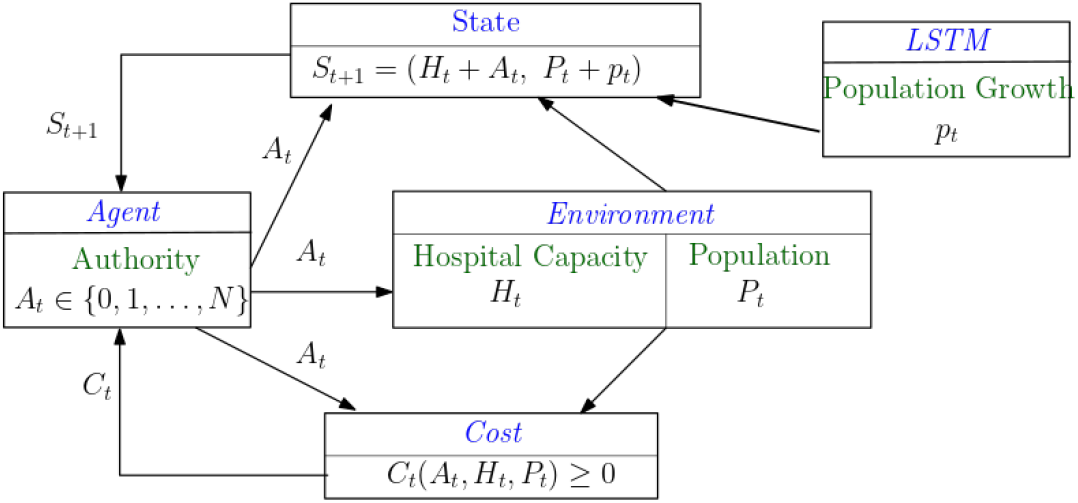
MDP Model for Policymaking [14].

1. *State S*_*t*_: Initial Population: *P*_0_ LSTM Population Growth Forecast: *p*_*t*_ Population at time step t: 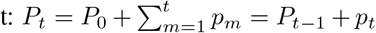. Initial Capacity: *H*_0_ Expansion Size(Action) at time t: *A*_*t*_ ∈ {0, 1, 2,, *N*} Capacity at time step t: 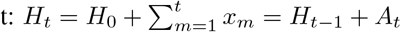
2. *Cost C*_*t*_: Daily Cost for *n*th day: Beds required : *r*_*n*_ ∼ Poisson Distribution(*mean* = *λ*(*P*_*t*_)^*a*^) Unattended Patients: *U*_*n*_=maximum of (*r*_*n*_ − *H*_*t*_) Hypothetical Cost for one unattended patient: *α* Cost: *c*_*n*_ = *α* × *U*_*n*_ Cost for *t*th time step: Number of Days in a time step: *N* Cost of adding 1 bed: *β* Augmentation cost for action *A*_*t*_: *C*_*aug*_ = *β* × *A*_*t*_ Total Cost: 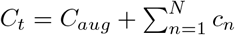
3. *Reward R*_*t*_: MDP agents aim to maximize their episodic cumulative reward. We assign a day to day costs *c*_*n*_ due to unattended patients and action cost *C*_*aug*_ for each time step. These costs are, in fact, negative rewards that the healthcare authority tries to minimize. So, the MDP reward for the agent is

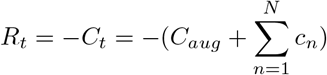 As reward maximization and cost minimization are equivalent in this problem; We elaborate on cost minimization for the rest of the report for easier understanding for the readers.
4. *Next State, S*_*t*+1_ : *S*_*t*+1_ = (*H*_*t*_ + *A*_*t*_, *P*_*t*_ + *p*_*t*_).

### B. LSTM network for population prediction, p_t_

The long short-term memory (LSTM) network is popular for time series data forecasting for tasks like machine translation, speech recognition, text prediction. We use this state-of-the-art LSTM network to predict the population of the next time step, a task similar to the time series data forecasting. The critical part of the forecasting is choosing appropriate hyperparameters and layers of the Neural Network. First, we select the previous 20-time steps data as input for the LSTM network. The regional healthcare authority of our country has a population history of more than 20 years of all the Upazila they serve. This sorts one of the drawbacks of the work in [14] and makes this work nationwide implementable. The Fig. 7 shows the general structure we use in our simulations.

**Fig. 7.**
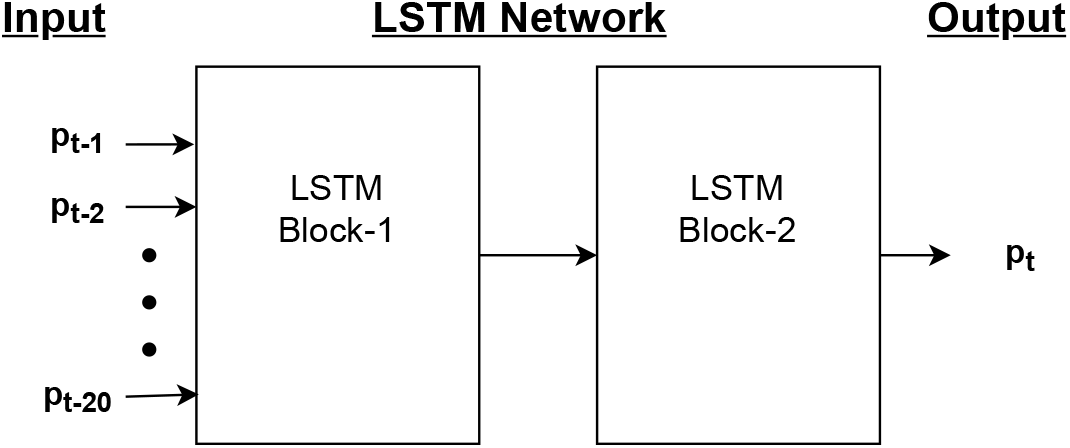
Neural Network Architecture for Population Growth Forecast.

### C. Optimal Policy

#### 1) Reinforcement Learning

The goal of the agent is to minimize following discounted cumulative cost in *T* time steps:

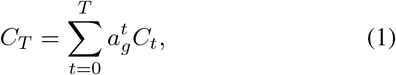

where *a*_*g*_ is the discount factor for future decisions. Cost in future carries less value than immediate cost and the discount factor resembles the weight of future cost for the agent. The agent’s objective is to minimize the long term cumulative reward providing, hence providing the optimal Long-Term capacity planning.

The solution lies in selecting optimal action *A*_*t*_ by solving the following Bellman equation, after *i*th iteration at time *t*, the agent’s value function is

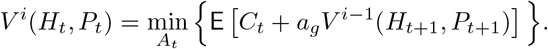

Since closed form solution is not tractable, the RL algorithm looks for the best action through simulations by assessing the expected cost E[*C*_*t*_] for each of the *t* + 1 possible actions. *C*_*t*_ is the immediate cost caused for the agent during time *t*. Since the agent’s action changes the next state of the hospital capacity, the future discounted cost through the value function of the next state, *V* ^*i−*1^(*S*_*t*+1_) depends on the action of the agent.

RL provides a very suitable framework to iteratively obtain the value function. The action-value function for our problem is:

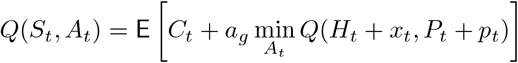

#### 2) Deep RL

The range of population growth for the region is extensive; hence the number of possible states is enormous. Thus, we require a neural network-based deep RL algorithm. The Advantage Actor-Critic (A2C) algorithm, a policy gradient-based algorithm, fits a continuous state environment. A2C applies two neural networks,

a. The actor-network, also known as the policy network, outputs probability for each action value through a softmax function. It aims to find the gradient of expected return *J* (*π*_*θ*_) of the policy *π*_*θ*_ with respect to the weights *θ* of the neural network by the following equation:

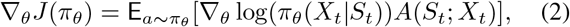

where the advantage function is given by

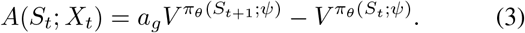
b. The critic-network is also identified as the value network to learn the value function for each state-action pair. In Eq. (3), 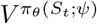 is the output of critic network for weight matrix *ψ*.

A pseudocode for the A2C algorithm is given in Fig. 8.

**Fig. 8.**
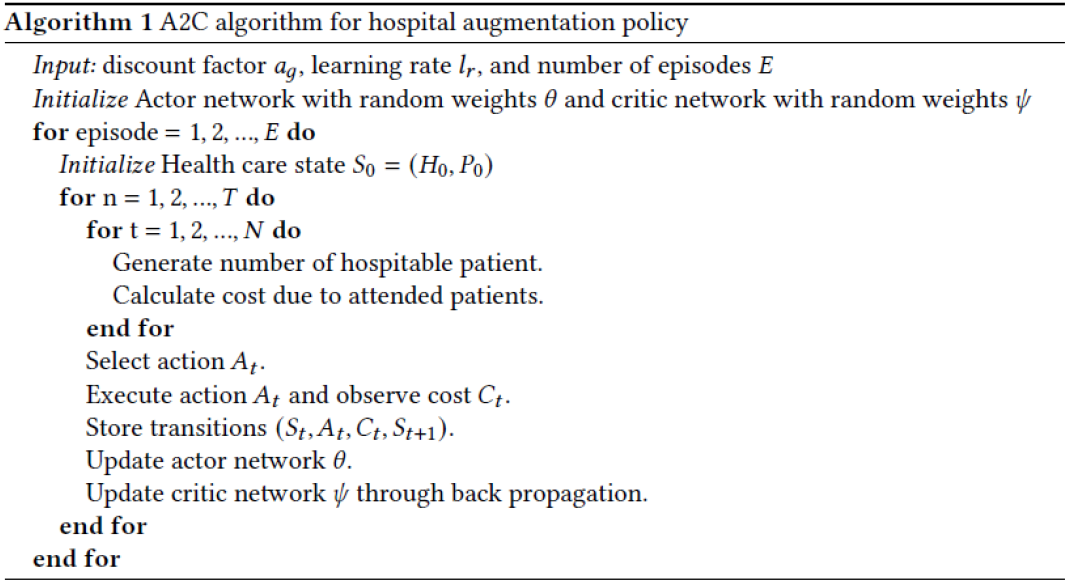
A2C algorithm for optimal policy [14].

## IV. Results

### A. Experimental Setup

#### 1) Zone Selection

We select Abhaynagar Upazila of Jessore district for the experiment. It has approximately 250 sq km area and a population of 232162 as of the year 2021 [20]. There are some private chambers of Physicians in the Upazila who provide health consultancy. However, the residents depend solely on the Government Upazila Health Complex as there is no private hospital to the best of our knowledge. The Abhaynagar Upazila Health Complex was established in 1978 with a current capacity of 50 [21]. So, we determine the two initial quantities *P*_0_ = 232162 and *H*_0_ = 50.

#### 2) Data Processing

The historical population data of Abhaynagar from [22] is the input to our LSTM network. We used the data between 1981-2001 for training and between 2001-2011 to validate our LSTM network as shown in Fig. 9. The LSTM network attains 99.37% accuracy in forecasting, which is impressive for a region’s population growth forecast.

**Fig. 9.**
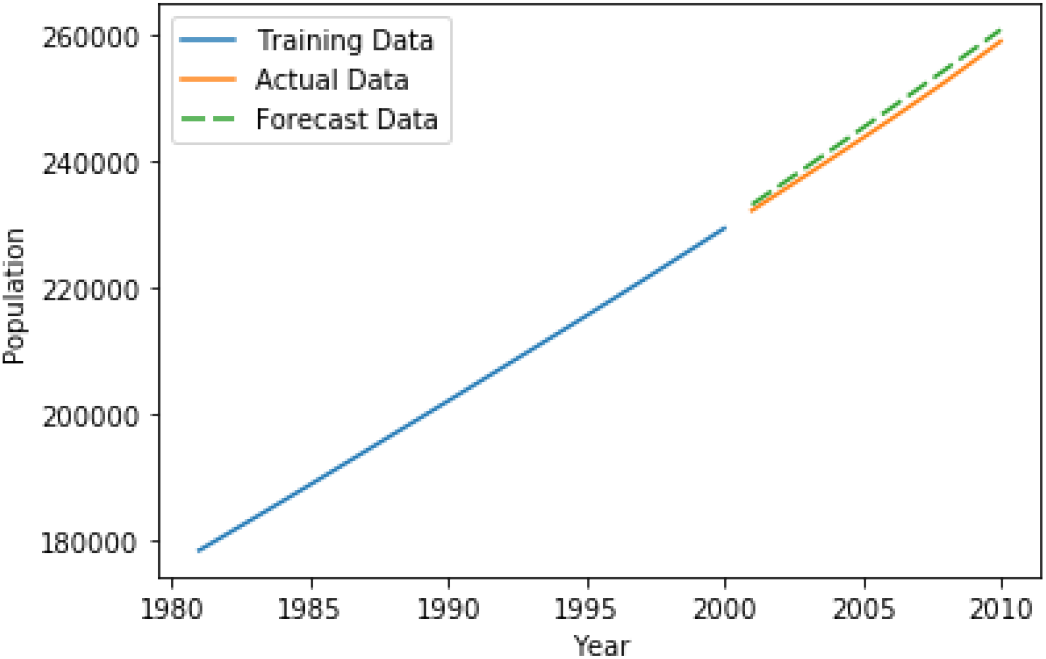
LSTM population prediction. Solid line represents the actual data and the dashed line represents the predicted population.

Furthermore, the LSTM network captures the trend fast, so if the Upazila experiences a major trend in its demography, the LSTM network catches the trend fast. The Hyperparameters for the LSTM networks are shown in Table I.

**Table I:**
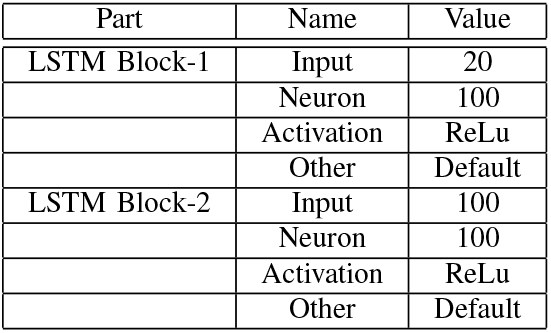
Hyperparameter of the LSTM Network.

For the cost analysis, we select Cost for one unattended patient, *α*= 50,000 taka and Cost of adding 1 bed, *β*=10,00,000 taka based on our survey of the local construction companies and residents of the Upazila.

### B. Comparative Analysis

Apart from the deep RL-based policy, we discuss two other myopic policies for a 30-year scheme.

#### 1) Rigid Policy

Often the healthcare authority ignores researching capacity expansion and follows fixed yearly expansion. We name this Rigid policy as it does not adapt to the scenario. Sometimes the central healthcare authority does not take recommendations from the local authority. So, this policy represents an unresponsive authority, however, a likely one for rural areas. This policy is expected to do worst, hence serving as the baseline to visualize other policies’ benefits. We do a grid search to find the optimal size to be 8 yearly inclusion of hospital beds that minimizes cost for this policy.

#### 2) Complaint-Based Policy

It is often difficult to determine the critical healthcare conditions for making capacity expansion decisions. The number of unattended patients for the previous time step for different Upazila can provide a reasonable guideline for the healthcare authority. They may use this number of denied patients as the decision-making criterion. The authority determines the optimal number proportionate to the number of complaints. In our simulation setup, we find that the optimal capacity expansion for this policy is,

Required augmentation size=(Number of Complaints from previous year)^*δ*^

Augmentation size= Minimum (Limit, Required augmentation size)

Here we set the Limit=10 as the maximum augmentation size. *δ* is the complain coefficient that we determine 0.6 to be optimal as it gave the minimum cost for a range of values between 0 to 1 for *δ*.

#### 3) Deep RL-Based Policy

We follow the actor and critic networks for the deep RL-based policy discussed in Project Design. We use a learning rate of 0.0001 and discount factor *γ* = 0.99 for the simulation. The proposed neural networks converge within 4000 episodes and learn the optimal policy as shown in Fig. 10.

**Fig. 10.**
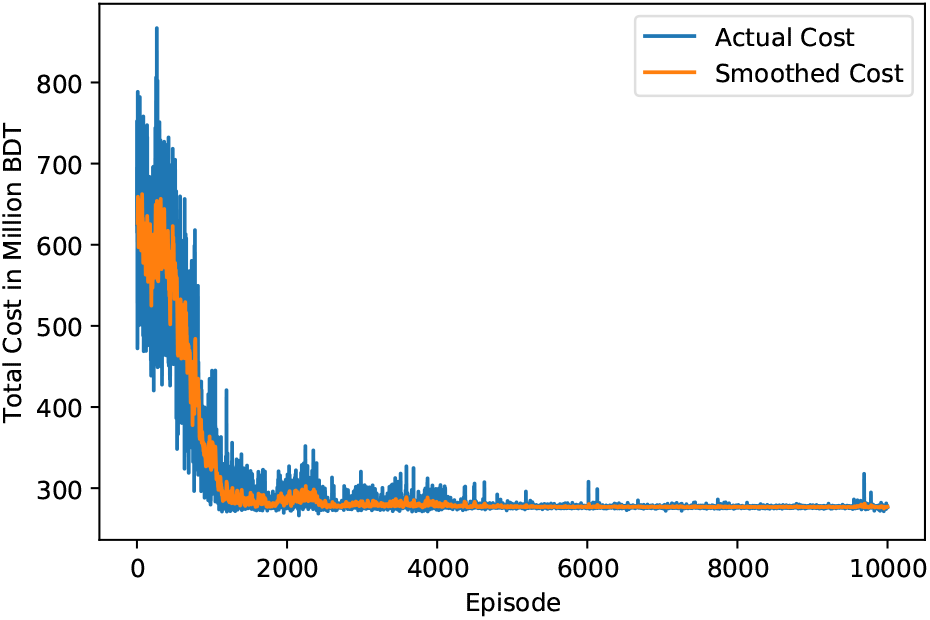
Cost convergence for the Deep RL based policy.

The Hyperparameter for the Deep RL network is shown in Table II.

**Table II:**
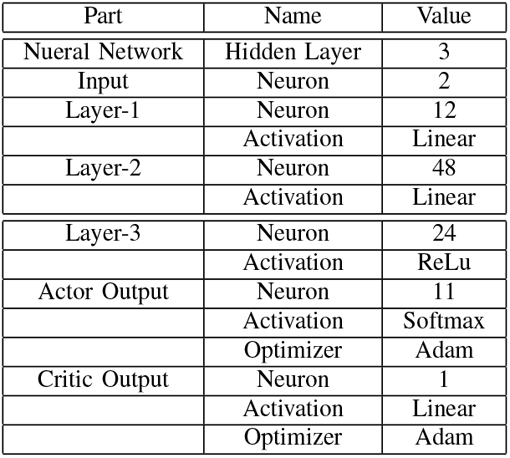
Hyperparameter of the Deep RL Network.

We compare the three policies stated above and find out that the deep RL-based policy outperforms the other two by a significant margin both in cost and complaint minimization. The result is summarized in Table III, where we take the complaint-based policy as a baseline for numerical comparisons, as this is the worst policy among the three.

**Table III:** Cumulative cost and complains over a thirty-year timeline.

| Policy | Cumulative cost (BDT) | Reduction | Complains | Reduction |
| --- | --- | --- | --- | --- |
| Rigid | 643.8 M | 0% | 17933 | 0% |
| Complain based | 490.2 M | 23.8% | 13833 | 22.9% |
| Deep RL based | 277.8 M | 56.8% | 6670 | 62.8% |

**Table IV:**
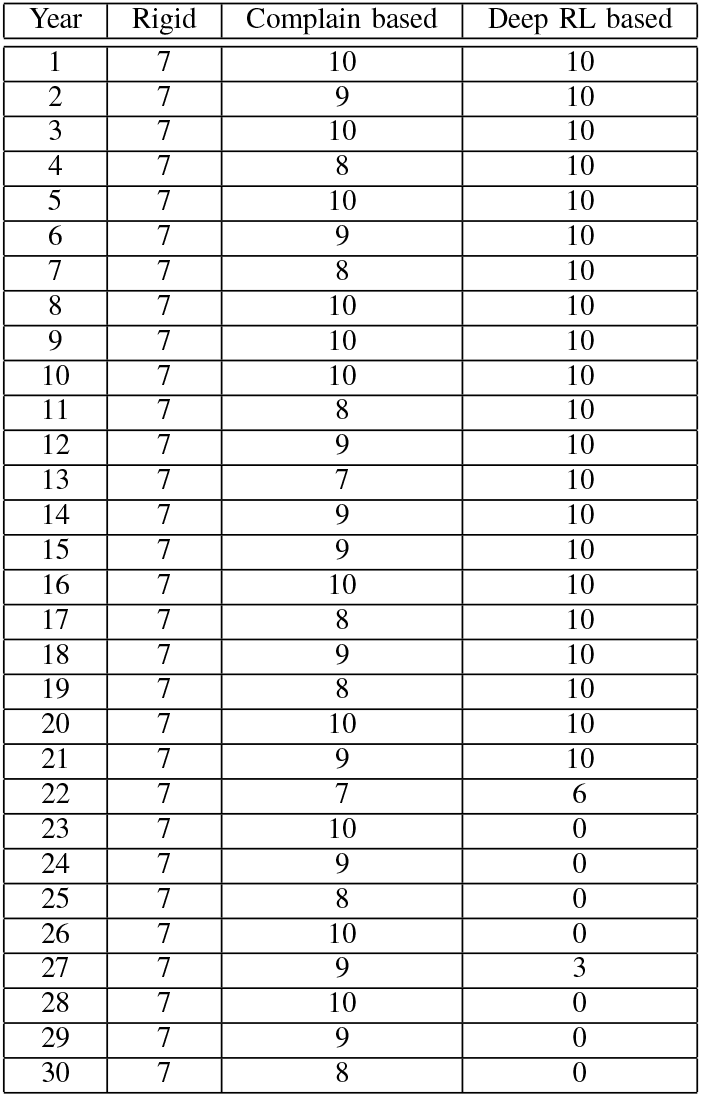
Yearly expansion under different policies.

| Year | Rigid | Complain based | Deep RL based |
| --- | --- | --- | --- |
| 1 | 7 | 10 | 10 |
| 2 | 7 | 9 | 10 |
| 3 | 7 | 10 | 10 |
| 4 | 7 | 8 | 10 |
| 5 | 7 | 10 | 10 |
| 6 | 7 | 9 | 10 |
| 7 | 7 | 8 | 10 |
| 8 | 7 | 10 | 10 |
| 9 | 7 | 10 | 10 |
| 10 | 7 | 10 | 10 |
| 11 | 7 | 8 | 10 |
| 12 | 7 | 9 | 10 |
| 13 | 7 | 7 | 10 |
| 14 | 7 | 9 | 10 |
| 15 | 7 | 9 | 10 |
| 16 | 7 | 10 | 10 |
| 17 | 7 | 8 | 10 |
| 18 | 7 | 9 | 10 |
| 19 | 7 | 8 | 10 |
| 20 | 7 | 10 | 10 |
| 21 | 7 | 9 | 10 |
| 22 | 7 | 7 | 6 |
| 23 | 7 | 10 | 0 |
| 24 | 7 | 9 | 0 |
| 25 | 7 | 8 | 0 |
| 26 | 7 | 10 | 0 |
| 27 | 7 | 9 | 3 |
| 28 | 7 | 10 | 0 |
| 29 | 7 | 9 | 0 |
| 30 | 7 | 8 | 0 |

The following table shows the number of bed allocation for each policy. The rigid policy makes fixed 7 augmentation each year. The complaint based policy reacts based on number of complains each year that varies with experience. However, the deep RL based policy correlates the number of beds with population and makes earlier investment to cope up with the need.

## V. Discussion

This work aims to provide hospital capacity expansion guidelines to the healthcare authority. We figure out several limitations to the approach that paves the way for further research. First, it is known that older people and children require more medical care than young adults. We generated hospital admission predictions based on the area’s total population, whereas age-based data might provide a more realistic scenario. Including some other features like occupation, weather data may increase the reliability of this model.

Furthermore, the model does not incorporate private medical facilities in the policy-making; including them may make this model implementable for urban areas. Finally, We use a discount factor *a*_*g*_= 0.99 for the case study, which resembles the healthcare is farsighted to give high weightage in future cost. This discount factor can be modeled based on the sincerity of the healthcare authority. For example, a selfish health care authority may try to minimize immediate costs by postponing capacity expansion work; that may get a discount factor *a*_*g*_ close to 0. So, future research may include directions to set the discount factor for different types of healthcare authority. Additionally, this setup may also be utilized for pandemic situation handling.

## VI. CONCLUSION

We have worked on the hospital management and cost minimization policy based on an MDP model in our paper. The case can be formulated as a MDP network where deep reinforcement learning determines the optimal value of bed up-gradation. LSTM network is used here for population prediction. Moreover, the model only takes population growth as the deciding factor, which makes it simple enough. Healthcare capacity expansion projects are expensive investments, and our goal is to minimize the cost. Existing literature provides general models, however we focus on the feasibility of the model for a specific area. Our experiments are based on Abhaynagar upazila of Jessore city, where the results show the benefit of this approach. So, this work has the potential to be studied for healthcare capacity expansion plans for our country.

## Data Availability

For the case study; we used population and hospital capacity data for Abhaynagar, jessore, Bangladesh from the following links.

http://203.112.218.65:8008/WebTestApplication/userfiles/Image/PopCenZilz2011/Zila_Jessore.pdf

http://facilityregistry.dghs.gov.bd/

https://en.banglapedia.org/index.php/Abhaynagar_Upazila

